# Feasibility of a high-dose behavioural exercise intervention on upper limb motor function in chronic stroke survivors

**DOI:** 10.1101/2023.07.31.23293284

**Authors:** Onno van der Groen, Jimena Garcia-Vega, Kirsten van Rijn, Dylan Edwards

**Affiliations:** School of Medical and Health Sciences, Edith Cowan University, Joondalup, Australia; Sir Charles Gairdner Hospital, North Metropolitan Health Service, Nedlands, Australia; Moss Rehabilitation Research Institute, Elkins Park, PA, United States

**Keywords:** Stroke, motor recovery, upper limb, neuroanimation, rehabilitation

## Abstract

**Background:** Stroke is a leading cause of adult disability and high-dose interventions may help reduce it. However, current practice does not allow for this.

**Purpose:** This study tests the feasibility of a high-dose upper limb therapy in chronic stroke survivors using a neuroanimation therapy (NAT).

**Methods:** Four chronic stroke survivors underwent 20 NAT sessions, 3 or 5 times a week for 90 minutes time-on-task. Feasibility was assessed with compliance to number of sessions and total time-on-task. Secondary outcomes included Fugle-Meyer Upper Extremity motor score (FM-UE), Action Research Arm Test (ARAT), grip strength, movement kinematics and cognition assessed using robotic technology.

**Results:** All participants attended the 20 prescribed sessions on the 3-day per week schedule. Two completed 90 minutes time-on-task in all sessions. Two showed clinical improvements in their FM-UE and ARAT. Movement kinematic analysis demonstrated improvements in motor control and cognition, however these changes did not seem to last when re-tested 1 month after the last training session.

**Conclusion:** 20 sessions of NAT is feasible in the chronic phase of stroke recovery with a 3-day per week schedule. Clinical improvements in arm function were observed in this high-dose upper limb NAT therapy, in one mild and one moderately affected stroke survivor.

## Introduction

More than 65% of stroke survivors experiencing lasting impairment to motor function [1]. Contrary to prior believes, significant changes in arm function can be possible with practice of suSensory-motor function can be improved in neurological patients in the chronic phase with practice of sufficient dose [2]. A Cochrane review found moderate evidence that upper limb (UL) function after stroke can be improved by providing at least 20 hours of additional repetitive UL task training[3], more recently, it was shown that 90 hours of therapy time in the chronic phase can lead to clinically important differences in measures of activity and impairment[4]. Multiple factors prevent most people from attaining this intervention dose. Some of these factors are 1) limited options for rehabilitation in the chronic phase; 2) not all rehabilitation is engaging and 3) the number of repetitions during standard (physiotherapy) practice is low[5]. The advent of technology that can enable immersive and engaging, high-dose practice, has led to new opportunities for restoring function in neurological patients [6]. In this study we used a neuroanimation therapy (NAT), consisting of motion capture and gaming system (MindPod, MSquare Health), which enables fun and engaging exploration of movement during task-specific practice. In this task patients engage the UL in gross and semi-fine movements to guide a dolphin to swim in a gaming environment. The system has been successfully used in subacute stroke where training led to similar results as a dose matched occupational therapy (OT) intervention [7], however feasibility in a chronic population has not been established yet.

## Methods

### Participants

Participants were recruited through local media outlets and through the ECU SPIN registry, between July 2020 and August 2022. Inclusion criteria were at least 18 years of age, a confirmed diagnosis of stroke, >6 months post stroke, commitment to attend all sessions and ability to follow simple instructions. Exclusion criteria were severe UL pain, fixed contraction, neglect, requiring manual physical assistance and aphasia. All participants gave informed consent for the study which was conducted in accordance with the ethical standards of the Declaration of Helsinki and received ethics approval from the ECU Human Research Ethics Committee (HREC ref. no. 2019-00873-VANDERGROEN). This study was registered with ANZCTR (trial Id: ACTRN12620001064998). At the baseline assessment, participant demographics were obtained, this demographics questionnaire was based on the stroke roundtable recommendations ([8]see table 2).

### Study design

We conducted a randomized feasibility trial where community-dwelling chronic stroke participants were allocated to either 3-days a week (distributed), or 5-days a week (massed) of training. The intervention consisted of 20 sessions (regardless of massed versus distributed group assignment), of 90 minutes time-on-task of NAT at the Robotics and Neurorehabilitation laboratory at the ECU Joondalup campus. In this intervention 3D movements of the paretic UL controlled a virtual dolphin, swimming through different underwater scenes with various task goals, including chasing and eating fish, eluding attacks and performing jumps (MSquare Health, a MindMaze Inc Company, see figure 1). Tasks were designed to promote movements in all planes and titrated based on successful completion of progressive levels of difficulty. Breaks were provided when necessary.

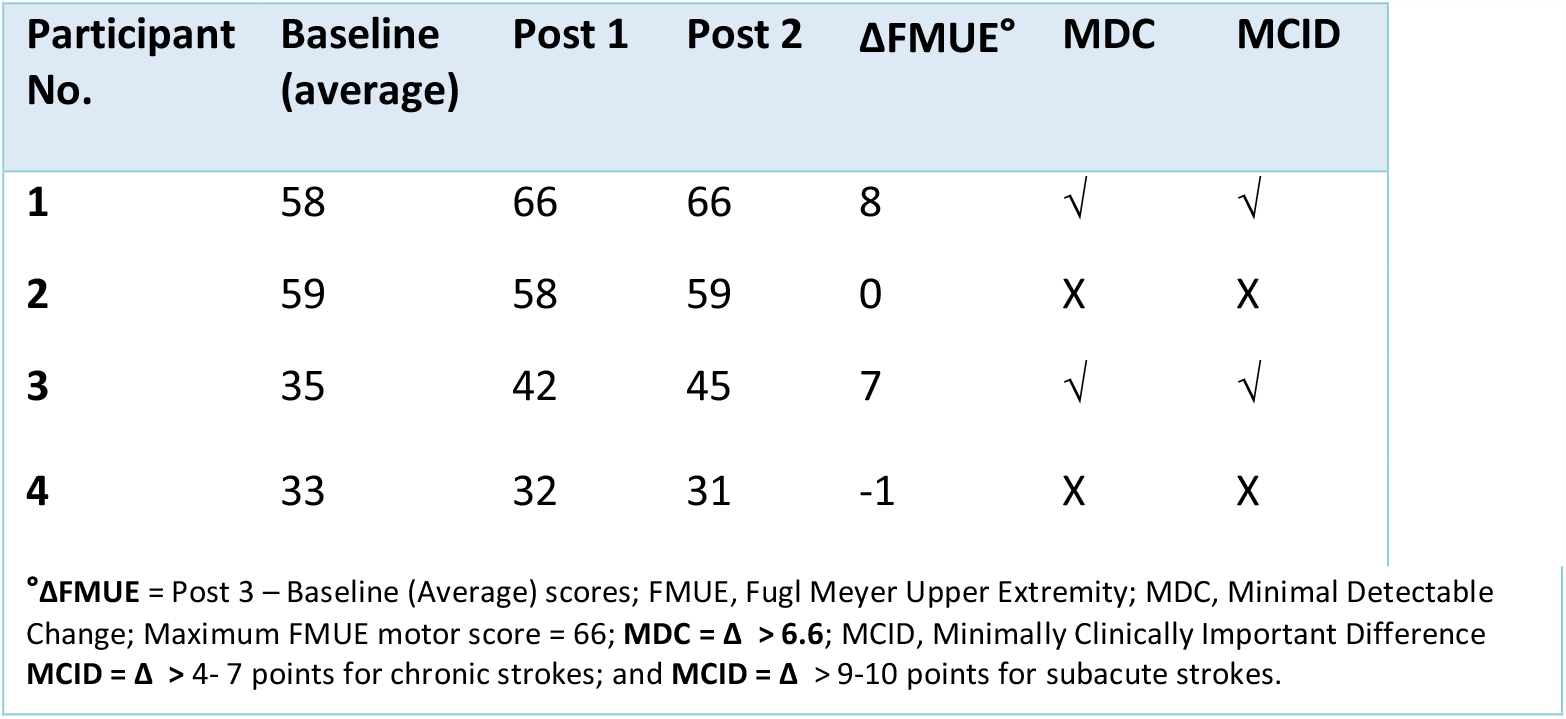

Participants wore an exoskeleton (EksoBionics) during training to provide anti-gravity support to their ULs. This exoskeleton has 5 different spring stiffnesses, with higher spring levels providing more support. The spring levels were adjusted during the intervention based on therapist’s recommendation and patient feedback. That is, spring support was increased when patients were not able to lift their arm up to 90 degrees of shoulder flexion. There were two baseline assessments within one week, where FMUE assessments were performed twice to ensure that patients were in a stable phase. If there were more than 2 points change in FMUE, a third FMUE assessment would be required. The post assessments were completed within 3 days, and a final assessment one month after the last training session.

### Outcome Measures

Assessments were completed at baseline (twice in the space of one week, pre1 and pre2), within 3 days of the last training sessions (post1) and 1 month after the last training session (post 2). The primary outcome of this study was feasibility.

Feasibility was assessed by participant’s compliance to attend 3 or 5 therapy sessions per week, and compliance / feasibility of participants being able to train for 90 minutes time-on-task. Adverse events were monitored in order to determine if there are any adverse reactions to this intervention.

A number of secondary outcomes were also assessed and are described in Table 1.

| Outcome Measure | Description |
| --- | --- |
| Physical Activity Enjoyment Scale (PACES) | An overall 18-item enjoyment for physical activity score to assess how much subject liked or disliked the intervention[9]. |
| Fugl-Meyer Upper Extremity scale (FMUE) | A stroke-specific, performance-based impairment index. It is designed to assess motor functioning, sensation and joint range of motion in patients with post-stroke hemiplegia[10]. |
| Action Research Arm Test (ARAT) | The ARAT is an evaluative measure to assess specific changes in upper limb |
|  | function in stroke patient with hemiplegia. It assesses a patient's ability to handle objects differing in size, weight and shape and therefore can be considered to be an arm-specific measure of activity limitation [11]. |
| Movement Kinematics and cognition | Quantitative assessment of movement will be performed using the KINARM exoskeleton robot (BKIN Technologies Ltd., Kingston, ON, Canada). The KINARM uses advanced robotics technology to quantify human voluntary motor control through flexion and extension movements at the shoulder and elbow joints. This robot has also been successfully used to assess cognitive function[12–15]. |
| Grip strength | Grip strength assessed with a handheld dynamometer |
| Quality of life (EQ-5D) | A standardised self-reported questionnaire to measure health status in stroke patients. The scale asks short tick box questions around mobility, personal care, usual activities, pain and anxiety[16]. |
| Depression Anxiety Stress Scale (DASS) | A standardised scale to assess anxiety, depression and stress symptomatology[17]. |
| Intrinsic Motivation Inventory (IMI) | This standardised scale will be used to measure participant's interest/enjoyment, perceived choice, and the value of the intervention[18]. |
| Modified Rankin Scale (mRS) | This scale assesses disability in patient who have suffered a stroke[19]. |

### Randomisation

Eligible participants were randomly assigned to receive either a massed or distributed practice schedule. When patients were not able to commit to 5 days a week, they were moved to the 3 days a week group. As this was a pilot feasibility study, the recruitment and data analysis were all carried out by the same research fellow who was not blind to the treatment allocation.

### Statistical analysis

The primary aim was feasibility; therefore data are represented in a descriptive manner as a case study series. Since this study was set up as a feasibility study, it was not adequately powered to run statistical analysis on group-level changes in outcome measures.

### Results

A total of 9 people expressed interest in this study. Two people were not eligible due to severe aphasia, three people did not respond after receiving the study information. A total of four participants enrolled and completed this study. At the first study visit participants completed a demographics questionnaire (see table 2).

**Table 2.**
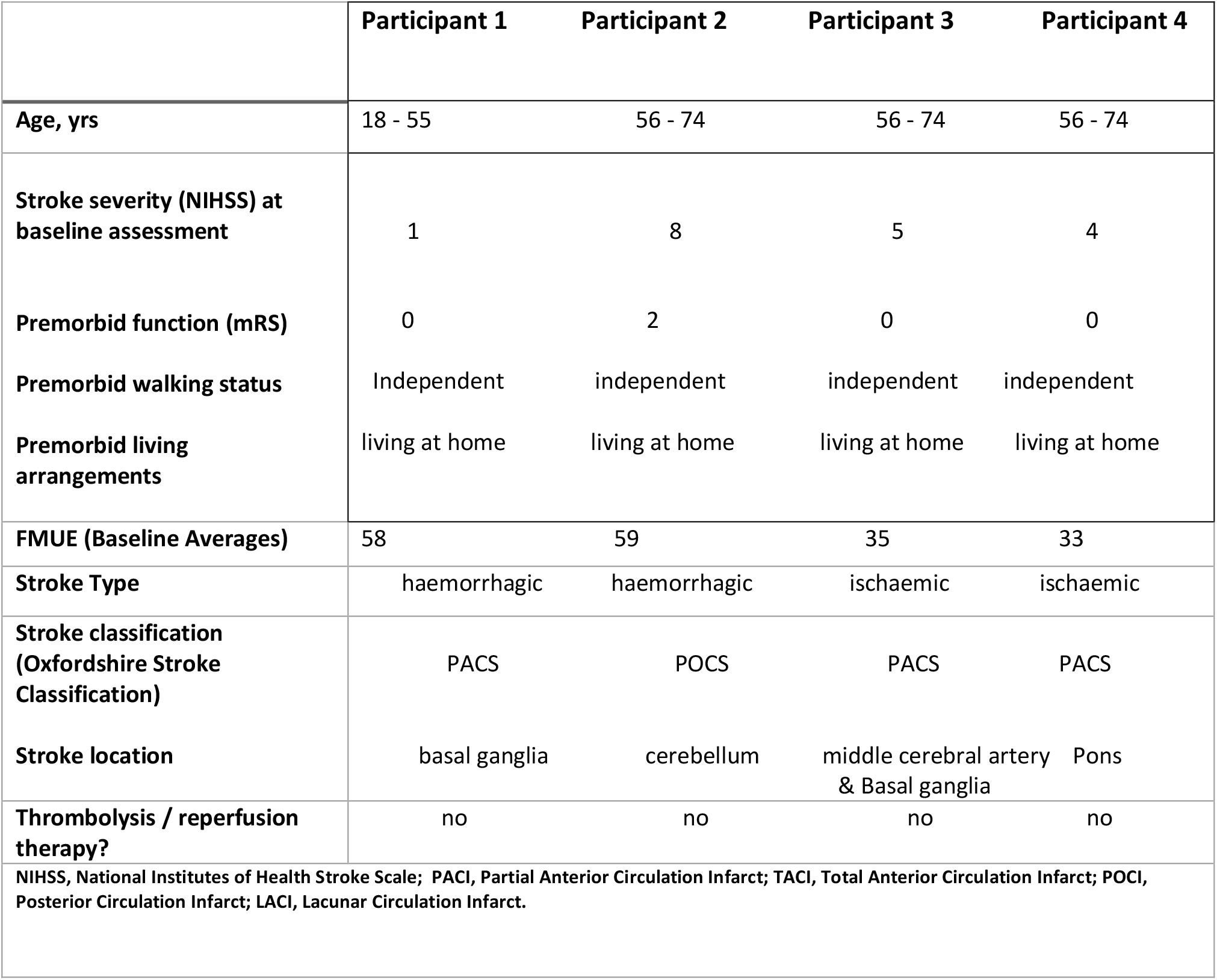
Baseline and demographic characteristics for pilot study participants [8].

#### Primary study aims: feasibility

All four participants were able to attend all their 20 training sessions. Participant 1 and 2 reached on average 90 and 92 minutes time-on-task. Participant 3 and 4 reached on average 65 and 69 minutes time-on-task. The total time-on-task from all 4 participants over 20 sessions was 29.9; 30.6; 21.7; 23 hours respectively. None of the participants were able to commit to five days-a-week of training. There was one adverse event (shoulder pain after a visist to a physiotherapist), which was not related to the study intervention.

#### Secondary study aims: outcome measures

Three of the participants indicated that they enjoyed the intervention (paces ranges from 18 – 126 with higher scores indicating more enjoyment, participants scored 117, 114, 69 and 126 respectively). FM-UE scores are shown in figure 1. Participant 1 (+9 points) and 3 (+7 points) demonstrated improvements in FM-UE larger than the minimal clinical important difference (MCID, [20]). Participant 2 and 4 did not improve their FM-UE. The ARAT showed a similar pattern where only participant 1 (+33) and 3 (+10) improved more than the MCID [21].

Movement kinematic assessments showed improved motor control in all participants for a range of parameters, that is irrespective of their clinical improvement. All participants demonstrated improvements in the path length ratio in a 2D-planar reaching task, that is the ratio of the length of the total movement relative to the length of a straight line between the initial position and target. This effect was only observed for the more affected (trained) arm and not for the less affected (untrained) arm, suggesting that this effect is due to the intervention. All participants, except participant 4 demonstrated smoother and faster movements of their paretic arm after the intervention. Arm spasticity was assessed with the elbow stretch test, where a flexion movement of the robot tests elbow extensors. All participants showed a decrease in the elbow flexion angle, that is the arm could not move as far in. This suggests that after the intervention the elbow extensors in the paretic arm became stiffer. Participant 1 and 4 showed a larger extension angle, suggesting that elbow flexors became less stiff. Cognitive functioning tests showed and improvement after the intervention on executive functioning (EF), assessed with the trail making B test. That is, all participants were able to perform the trail B faster, however 1 month after the intervention participants became slower but performance was better compared to baseline. A measure of working memory, assessed with spatial span, showed an improvement in all participants, however 1 month after the intervention the performance of participants 1, 2 and 4 decreased again, but performance was better compared to baseline. All participants showed an increase in hand grip strength in their more affected/trained hand (1.6%, 20.4%, 103%, 63.7% respectively). Only participant 1 still showed this increased grip strength at the one month follow up. All participants demonstrated normal levels of depression, anxiety and stress throughout the study. The intrinsic motivation inventory (IMI) showed that the score for value and enjoyment for participant 3 decreased after the intervention. Modified rankin scale stayed stable for participant 2 and 4. Participant 1 showed a decrease in mRS from 3 to 1, participant 3 showed an increase in mRS, from 2 to 3.

## Discussion

Our feasibility study shows for the first time that 20 sessions of NAT (comprising up to 90min time-on-task per session / 30 hours total) is feasible and that this could lead to clinically important changes in arm function, assessed with the FM-UE and ARAT. None of the participants were able to commit to 5 sessions a week, therefore it is not feasible to run this intervention for 5 days a week. Two out of the four participants managed 90min time-on-task, which were the least severely affected patients. The other two patients still managed 60min time-on-task, and a clinically relevant change in arm function was observed in one of these participants. This suggests that in the chronic, stable phase after stroke, improvements can be made with sufficient dose, and that this can also occur in moderately affected patients. Other studies have also demonstrated that high-dose therapy with technological interventions could improve arm function[25–28] and report similar benefits produced by a matched dose conventional therapy. An increase in the amount of training is crucial to drive neurophysiological changes and even in the chronic stage improvements can be made. Our study demonstrated that 3 of the 4 participants enjoyed the intervention. The same participant who did not enjoy the intervention showed a reduction in the intrinsic motivation inventory. This participant was more severly affected (baseline FM-UE of 35), therefore the intervention could have been too challenging for the participant. Enjoyment is a positive emotion linked to intrinsic motivation, meaning that a behaviour is performed for the enjoyment it provides[22]. Enjoyment is a predictor of physical activity level in healthy elderly adults[23]. Moreover, a previous study in chronic stroke survivors demonstrated that enjoyment was greatest for a custom video game, compared to standard care and an off-the-shelf video game[24].

Movement kinematic data shows improvements in a range of parameters, irrespective of clinical change. These movement kinematics measure the quality of the movement, and allow to distinguish changes associated with behavioural restitution from compensation[29]. This indicates that (i) our intervention can have a positive influence on the quality of the movement, and (ii) even in patients that do not show changes on a clinical scale, the intervention could improve movement quality. The kinematic analysis was performed in the 2D space, therefore participants were not required to lift their arm against gravity, which they did have to do in the clinical analysis.

Measures of executive functioning and working memory improved directly after the 20 sessions. We only recorded one adverse event, not related to the intervention. This implies that a high-dose intervention can be safely administered.

This study have several weaknesses. This study had the limited sample size, a lack of a control group and relative low recruitment rate. Whether this intervention results in better outcomes than dose matched regular therapy is unknown. Other studies suggest that it might not be the case [25–28]. Possible explanations for the low recruitment rate are the large time commitment, the travel involved and financial burden for transport to the study site. Therefore, this intervention might be more suitable in an outpatient hospital setting.

## Conclusion

High-dose upper-limb rehabilitation training in the chronic phase is feasible with a 3-day per week schedule. Less severely affected patients are able to tolerate 90 minutes time on task per session. The outpatient setting of this study is not the ideal place for this intervention.

Future studies could make this intervention available as a telerehabilitation intervention or as part of the hospital outpatient service such that treatment frequency could be increased.

## Data Availability

All data produced in the present study are available upon reasonable request to the authors.

